# A placebo-controlled clinical study to assess the impact of variable complex weak magnetic fields (VCMF’s) generated by the BeCurie™ (Evolv28) device on the subjects with perceived stress and anxiety

**DOI:** 10.1101/2023.08.03.23293534

**Authors:** Mohan Krishna Jonnalagadda, Lalitha Palle, Shyam Sunder Pasumarthi, Chaitanya Chakravarthi Gali

## Abstract

**Background:** Exposure to variable weak magnetic fields, reported to have shown beneficial effects on several neurological illnesses. However, existing therapies are costly, complex, and lack subject ease for frequent follow ups. In the current study, the novel BeCurie™ (Evolv28) wearable neck device that emits variable complex weak magnetic fields (VCMF’s) is evaluated for its positive impact on subjects with perceived stress and anxiety.

**Methods:** Eighteen participants were enrolled in this study. The primary outcomes of the study were to assess the improvements in perceived stress and anxiety symptoms in the BeCurie™ treated group. Stress and anxiety scores were assessed using DASS-21, HAM-A, and PSS. Quality of life was assessed using the MQoL-R questionnaire. Serum Cortisol and complete blood profile were assessed to understand the safety profile of BeCurie™ treatment.

**Results:** Participants in the BeCurie™ group showed a significant reduction in stress and anxiety scores compared to the placebo group on Day 30. Furthermore, open label study assessments on Days 60 and 90 revealed improvements in self-reported stress and anxiety scores, significant time dependent improvements in all major domains of quality of life, including physical, psychological, existential, and support-based aspects of life. No adverse events were reported during the study. Comprehensive blood profile assessment showed no significant changes in either the placebo or BeCurie™ groups.

**Conclusions:** The findings indicate that VCMF’s emitted by the BeCurie™ device can be a supporting non-invasive alternative therapy for managing stress and anxiety. Nonetheless, the limitations of the study, including the small sample size and the lack of a follow-up assessment beyond 90 days, suggest that further investigations are needed to establish the long-term efficacy of BeCurie™ in managing stress and anxiety symptoms.

## Introduction

In 2020, the World Health Organization (WHO) reported that one in eight adults, or 970 million individuals worldwide, experienced mental health disorders^1^. The COVID-19 pandemic has led to the emergence of several mental health issues, including anxiety, sleeplessness, stress, depression, post-traumatic stress syndrome, and other psychological problems due to physical, societal, and economic impacts^2^. According to WHO reports as of March 2022, the COVID-19 pandemic has triggered a 25% increase in the prevalence of anxiety and depression worldwide^3^.

Magnetic fields are ubiquitous in our daily life, with varying degrees of exposure depending on the location and occupation^4, 5^. These fields may arise from natural sources such as the geomagnetic field, intense solar activity, and thunderstorms, as well as human-made sources like factories, transmission lines, electric appliances, magnetic resonance imaging, and medical treatments^6^. It is well-established that weak magnetic fields can modulate molecular and cellular responses, complex physiological processes, including activation of certain biological pathways like nitric oxide, induce mitophagy, sympathetic system, and further impact even behavioral and mood changes^7–11^. Additionally, extremely low frequency weak magnetic field (ELF-MF) therapy has proven efficacious in supporting depressive disorders^12, 13^, shown to improve serotonin levels^14^, moderate blood pressure^15, 16^ and can serve as an alternative therapy for drug-resistant or post-stroke patients^17^.

Several human clinical studies have reported the beneficial effects of short and long-term exposure to pulsated and complex magnetic fields (in the order of picotesla (pT) and microtesla (µT)) in subjects with various neurological illnesses. Nishimura et al. demonstrated that repeated exposure to 1 µT extremely low frequency magnetic fields can benefit people with hypertension and helped with reducing their dependence on hypertensive medication^15^, Baker-Price and Persinger (1996, 2003) found that applying a weak, complex burst-firing magnetic field once every three seconds across the temporal lobes on a weekly basis was linked to a significant decrease in both psychometric depression and clinical depression in patients^18^, According to Anninos et al., administering pico Tesla - transcranial magnetic stimulation (pT-TMS) to each functional point of seizure activity led to a decrease in emitted power from the affected area and a reduction in epileptic activity^19^, Applying pT-TMS to children with autism disorder was found to impact their beta rhythm, resulting in an increase towards the frequency range of 18-26 Hz^20^. In a small pilot study, pT-TMS at low frequencies (≤1 Hz) had a beneficial effect on individuals with Alzheimer’s disease^21^, Multiple Sclerosis patients who underwent pT-TMS treatment at home for one-month experienced treatment benefits^22^, In another study, it has been demonstrated that, pT-TMS at a frequency of 2 Hz and intensity of 7.5 pT for 6 minutes resulted in a rapid and substantial reduction in Parkinson’s disease disability and a near-complete resolution of the dyskinesis ^23, 24^. Anninos et al. demonstrated the efficacy of pT-TMS in a small sample of individuals with cerebral palsy. The study revealed increased amplitudes across the 2-7 Hz range and substantial improvement and normalization of MEG recordings with the use of pT-TMS^25^.

Based on the current evidence, we have designed a novel wearable device (BeCurie™) that emits variable complex weak magnetic fields (VCMF’s) equivalent to magnetic flux density ranging between 0.4 to 10 milli gauss (0.04 to 1 µT) generated with a specific amplitude, frequency, phase, and rhythm. BeCurie™ is classified as a General Wellness - Low risk device by FDA (docket no: FDA-2014-N- 1039) and is compliant with CE (Conformité Européenne), SAR (Specific Absorption Rate), ISED (Innovation, Science and Economic Development Canada) and FCC (Federal Communications Commission) regulations with respect to safety and human exposure of electromagnetic fields generated by the device. In the current randomized double-blind pilot study, we evaluated the positive impact of the BeCurie™ device on subjects with perceived stress and anxiety.

## Methods

### Study Participants

Eighteen adults, encompassing both male and female individuals, aged between 18 to 60 years, were recruited for the study at Yashoda Hospitals located on Raj Bhavan Road, Somajiguda, Hyderabad, Telangana – 500082. The subjects had sought medical assistance for complaints related to self-perceived stress and anxiety, and as part of the primary screening process, subjects were asked to complete the DASS-21 assessment, with the guidance of a consulting physician. Participants who met the predefined eligibility criteria based on the primary assessment were then included in the study. Exclusion criteria encompassed a history of alcohol abuse, skin-related disorders, use of antipsychotic medications, pregnancy or lactation, presence of surgical implants such as pacemakers or defibrillators, inability to comprehend the procedures of the study due to language barriers, psychological disorders, or dementia, a serious medical condition such as head and neck cancer or skin cancer, as well as individuals with a history of allergy or hypersensitivity to any medical device or its components. The study was approved by the Institutional Ethics Committee Yashoda Academy of Medical Education and Research (IEC-YAMER) with registration number ECR/49/Inst/AP/2013/RR-19, while the clinical trial was recorded in the CTRI database under the study number CTRI/2022/03/041445. The study flow diagram is outlined in Figure 1.

**Figure 1:**
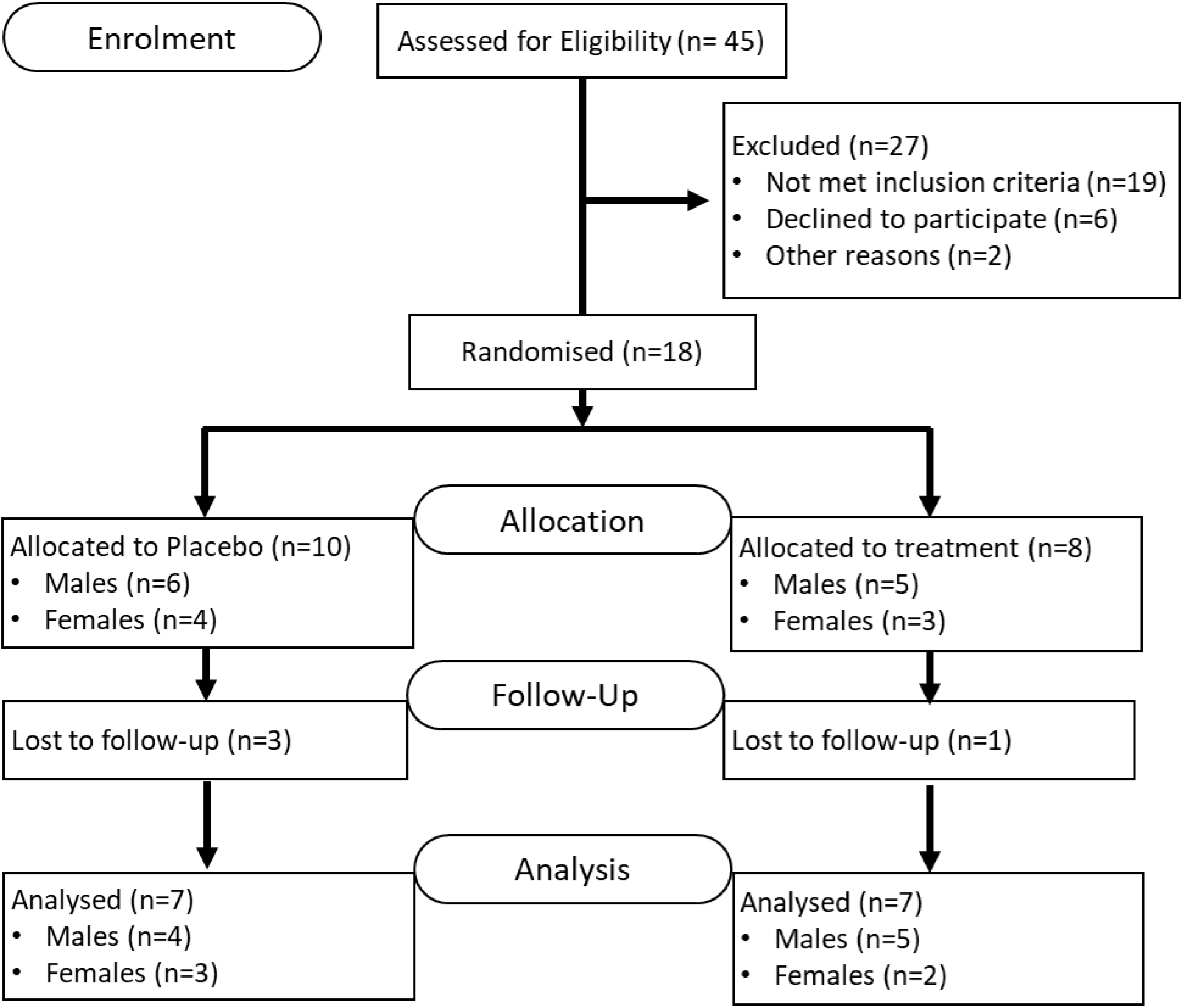
CONSORT diagram of participant flow.

### Experimental Design

This study utilized a randomized placebo-controlled parallel design, with participants undergoing preliminary screening that included medical inquiry, health screening, and physical examination upon enrolment. Only those with moderate to severe stress and anxiety were included. Participants were randomly assigned using block randomisation into two groups: the Placebo Group or Treatment Group. Prior to receiving the designated interventions, all participants signed the Informed Consent Form (ICF) and underwent measurements for baseline values. The study consisted of a 12-week design that included two stages: the double-blind phase (weeks 1-4) and the open-label phase (weeks 5-12). During the 1– 4-week period, both groups received their respective interventions. The treatment group received a functional device that emits VCMF’s, while the placebo group received a dummy device that operated but does not emit VCMF’s. Participants were advised to wear the device for a minimum duration of 8 hours per day and were advised to not to exceed the maximum duration of 10 hours of usage per day.

### Interventional device

Evolv28 or BeCurie™ (Aether Mindtech Pvt Ltd, Hyderabad) is a novel, compact wearable device that generates VCMF’s from the media positioned on the neck of the user (Figure 2). The VCMF’s are generated by passing variable long wave digital signals in an encrypted format passing through DAC, and then regulating and passing it through magnesium and zinc ferrite inductors placed along the neck. The digital signals are specifically designed with variable amplitudes, frequencies and phases within the range of 1Hz to 900Hz and sampling rate of 8 to 16 bit with variable range of 44KHz to 88KHz. These programs are controlled through a mobile application, ultimately producing the variable magnetic field with flux density ranging from 0.4 to 10 milli gauss (mG).

**Figure 2:**
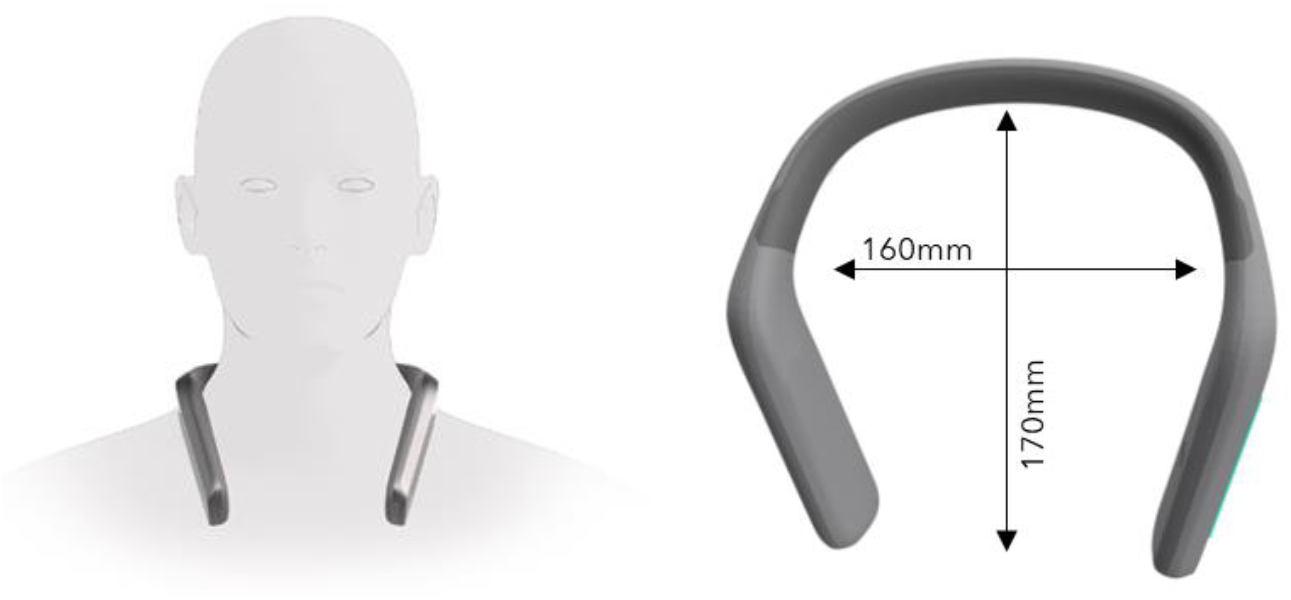
Placement of the BeCurie™ device (neck position). The device is positioned on the neck and rested on the shoulders of the user in such a way that it emits the VCMF’s form the media placed in the close proximity to user’s neck.

### Primary Outcome Measurements

The primary objective of this study was to evaluate the efficacy of VCMF’S intervention on perceived stress and anxiety levels in adults with moderate to severe symptoms. To measure the effectiveness of the intervention, objective and self-rated scales for depression, anxiety, stress, perceived stress, and quality of life were used. The study utilized the Depression Anxiety and Stress Scales - 21 (DASS-21) and the Perceived Stress Scale (PSS) to assess stress scores. The Hamilton Anxiety Index was used to evaluate anxiety, and the McGill Quality of Life (MQoL) scale was used to assess the impact of the intervention on the overall quality of life. Permissions for using the scales were acquired from the respective agencies.

The DASS-21 subscales for depression, anxiety, and stress each comprised seven items graded on a four-point scale ranging from 0 to 3. The depression subscale assessed dysphoria, hopelessness, and life devaluation, while the anxiety subscale assessed autonomic arousal, skeletal musculature symptoms, and situational anxiety. The stress subscale was sensitive to chronic nonspecific arousal and assessed difficulty relaxing, nervous arousal, being easily upset/agitated, irritable/over-reactive, and impatient. To produce equivalent scores to the full version of DASS (42-item), the total score of each subscale was multiplied by 2, resulting in scores ranging from 0 to 42. The cut-off scores for the DASS-21 subscales were as follows: normal (0 to 9 for depression, 0 to 7 for anxiety, and 0 to 14 for stress), mild (10 to 13 for depression, 8 to 9 for anxiety, and 15 to 18 for stress), moderate (14 to 20 for depression, 10 to 14 for anxiety, and 19 to 25 for stress), severe (21 to 27 for depression, 15 to 19 for anxiety, and 26 to 33 for stress), and extremely severe (28 or more for depression, 20 or more for anxiety, and 34 or more for stress)^26, 27^.

The Hamilton Anxiety Index (HAM-A) questionnaire comprises 14 components that encompass both psychological and physical symptoms. These components include anxious mood, tension (including startle response, fatigability, and restlessness), fears (including of the dark, strangers, and crowds), insomnia, cognitive symptoms (such as poor memory and difficulty concentrating), depressed mood (including anhedonia), somatic symptoms (including aches and pains, stiffness, and bruxism), sensory symptoms (such as tinnitus and blurred vision), cardiovascular symptoms (such as tachycardia and palpitations), respiratory symptoms (such as chest tightness and choking), gastrointestinal symptoms (such as symptoms of irritable bowel syndrome), genitourinary symptoms (such as frequent urination and loss of libido), autonomic symptoms (such as dry mouth and tension headache), and observed behaviour during the interview (restless, fidgety, etc.). Each item is scored on a simple numeric scale ranging from 0 (not present) to 4 (severe). Scores of >17/56 indicate mild anxiety, while scores of 25– 30 indicate moderate to severe anxiety^28^.

The Perceived Stress Scale (PSS-10) is a self-report questionnaire consisting of 10 items that measure an individual’s overall perceived stress. The scale produces a total score ranging from 0 to 40, with higher scores indicating greater levels of perceived stress. To calculate subscale scores, Factor 1 (“Negative”) is determined by adding the six negatively worded items (Items 1, 2, 3, 6, 9, and 10), while Factor 2 (“Positive”) is determined by adding the four positively worded items (Items 4, 5, 7, and 8). Higher scores on Factor 1 reflect stronger negative distress and stress sensations, while higher scores on Factor 2 indicate greater coping capacities. To obtain the total score, the four positively phrased items are reverse scored and then added to all of the scale items^29, 30^.

The McGill Quality of Life Questionnaire (MQoL) was originally developed in Canada for patients with life-threatening illnesses to measure their quality of life (QoL). The MQoL considers both positive and negative factors that contribute to QoL and assesses four areas: existential, social, psychological, and physical. It also includes a question to rate total QOL. The MQoL includes 16 items, with each item having a numeric response scale ranging from 0 to 10, with verbal responses at both ends. The score of 0 indicates the worst case after reversing scored items. For instance, “Over the past 48 hours, I felt physically dreadful (0) vs. physically well (10)” is one of the items. The MQoL questionnaire efficiently measures QOL while considering the length of the instrument^31^.

### Secondary Outcome Measurements

Blood cortisol assessment was conducted by collecting blood samples from each subject on two different days - Day 0 and 30 - between 3-4 PM. The collected samples were then processed to prepare serum aliquots, which were subsequently stored at a temperature of -20°C until further evaluation. Prior to analysis, the serum was thawed to room temperature. Each sample was analyzed twice to ensure accuracy of results. Serum cortisol levels were measured using a commercially available enzyme-linked immunosorbent assay (ELISA) kit (R&D Systems, Minneapolis, MN, USA), according to the manufacturer’s instructions.

### Safety Assessment

To ensure the safety of the participants, adverse events were monitored throughout the study. Additionally, comprehensive blood analysis was conducted during enrolment and at the first follow-up visit on Day 30. The analysis included two components: haematology and biochemistry. Haematology comprised a range of measurements, including haemoglobin, packed cell volume, red blood cells, mean corpuscular haemoglobin concentration, mean corpuscular volume, mean corpuscular haemoglobin, total white blood cells, differential count, complete urine examination, erythrocyte sedimentation rate, and platelet count. Biochemistry encompassed HbA1C, creatinine, serum lipid profile, total cholesterol, HDL cholesterol, LDL cholesterol, VLDL cholesterol, triglycerides, non-HDL cholesterol, cardiac risk ratio, serum calcium, urea, and uric acid, as well as liver function tests, total bilirubin, bilirubin conjugated and unconjugated, alkaline phosphatase, SGOT, SGPT, total protein, and albumin-globulin ratio. The analysis was conducted to identify any potential risks or adverse effects associated with device use.

### Statistical Analyses

Descriptive statistics were reported using means and standard deviations to summarize the changes observed. A paired t-test was employed to examine the impact of the intervention within the group, given the small sample size. To assess the effect of the intervention between the placebo and treatment groups, an unpaired t-test was conducted. Furthermore, a repeated-measures analysis of variance (ANOVA) was performed to evaluate the time-varying covariates of each outcome measurement in the treatment group. A p-value of 0.05 was considered as the threshold for statistical significance.

## Results

### Subject Demographics

In the conducted study, both male and female subjects were enrolled with a mean age of 41.85±8.31 and 42.42±11.50 years for the placebo and BeCurie™ groups, respectively, based on block randomization. The gender distribution was similar between the two groups, six males and four females were allocated to the placebo group, five males and three females were allocated to the BeCurie™ group. A total of eighteen subjects were enrolled for the study, but three subjects (three from the placebo group and one from the BeCurie™ group) failed to complete the follow-up assessments (Figure 1). The subjects were enrolled based on DASS-21 eligibility criteria, and individuals with moderate to extremely severe levels of stress and anxiety were included in the study. The baseline characteristics of both placebo and treatment groups were provided in Table 1.

**Table 1:**
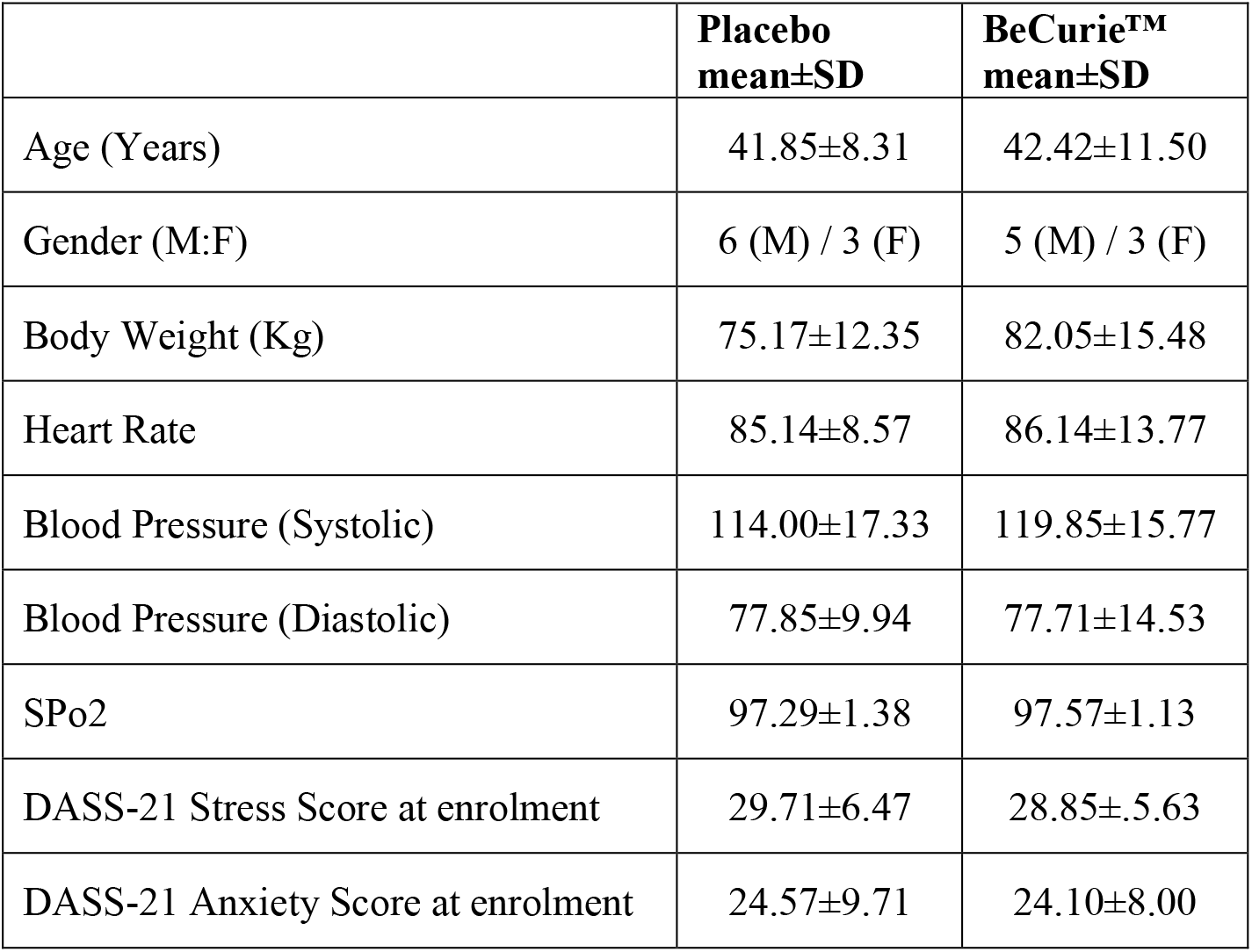
Demographic and clinical characteristics of the study population by treatment arm: ^a^Overall there are no significant differences between the arms.

|  | **Placebo**<br>**mean±SD** | **BeCurie™**<br>**mean±SD** |
| --- | --- | --- |
| Age (Years) | 41.85±8.31 | 42.42±11.50 |
| Gender (M:F) | 6 (M) / 3 (F) | 5 (M) / 3 (F) |
| Body Weight (Kg) | 75.17±12.35 | 82.05±15.48 |
| Heart Rate | 85.14±8.57 | 86.14±13.77 |
| Blood Pressure (Systolic) | 114.00±17.33 | 119.85±15.77 |
| Blood Pressure (Diastolic) | 77.85±9.94 | 77.71±14.53 |
| SPo2 | 97.29±1.38 | 97.57±1.13 |
| DASS-21 Stress Score at enrolment | 29.71±6.47 | 28.85±5.63 |
| DASS-21 Anxiety Score at enrolment | 24.57±9.71 | 24.10±8.00 |

### Assessment of Changes in Self-Reported Stress, Anxiety, and Depression Scores at Day 30

The primary outcomes of the study were improvements in stress and anxiety symptoms in the BeCurie™ administered group. The results indicate that the BeCurie™ group exhibited a significant improvement in stress and anxiety scores on day 30 (follow-up 1) compared to the placebo administered group. Specifically, the stress score as measured by DASS-21 (95% CI = 8.6 (0.22, 17.49), p=0.045) and PSS (95% CI = 8.1 (0.89, 17.1), p=0.032) were significantly lower in the BeCurie™ group. Furthermore, anxiety scores as measured by HAM-A (95% CI=11.9 (0.06, 23.93), p=0.049) were significantly lower in the BeCurie™ group compared to the placebo group (Table 2, Figure 3).

**Figure 3:**
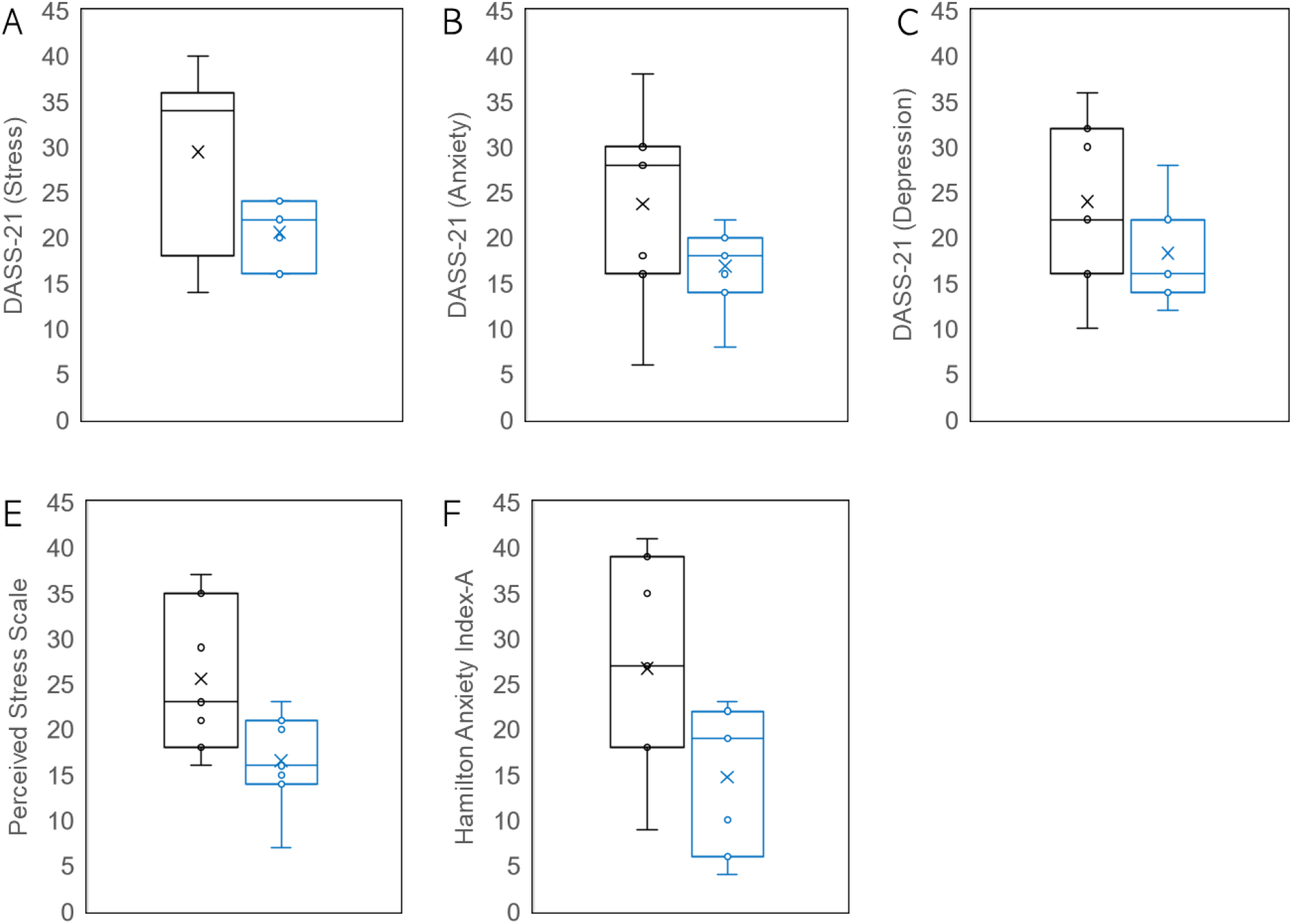
Assessment of stress, anxiety, and depression scores in Placebo (n=7) and BeCurie™ (n=7) groups on Day 30. The data is represented in a box chart showing the distribution of data, highlighting the mean and outliers.

**Table 2:** Assessment of stress, anxiety, and depression scores in Placebo (n=7) and BeCurie™ (n=7) groups on Day 30. Values are represented as mean ± SD. *P<0.05 was considered significant. Abbreviations: DASS-21 = The Depression, Anxiety and Stress Scale - 21 Items; PSS = Perceived Stress Scale; HAM-A = The Hamilton Anxiety Rating Scale.

| **Assessment** | **Placebo**<br>**(mean±SD)** | **BeCurie™**<br>**(mean±SD)** | **t-test** | **95% CI** | **p value** |
| --- | --- | --- | --- | --- | --- |
| DASS-21<br>(Stress) | 29.4±9.91 | 20.5±3.40 | 2.24 | 8.6 (0.22, 17.49) | 0.045* |
| DASS-21<br>(Anxiety) | 23.7±10.85 | 16.8±4.74 | 1.53 | 9.7 (-2.9, 16.61) | 0.152 |
| DASS-21<br>(Depression) | 24.1±9.23 | 18.2±5.82 | 1.38 | 8.9 (-3.27, 14.70) | 0.191 |
| PSS | 25.5±8.24 | 16.5±5.38 | 2.42 | 8.1 (0.89, 17.1) | 0.032* |
| HAM-A | 26.7±12.17 | 14.7±7.86 | 2.19 | 11.9 (0.06, 23.93) | 0.049* |

### Assessment of Changes in Self-Reported Stress, Anxiety, and Depression Scores over Time

Subsequent evaluations conducted on Days 30, 60, and 90 revealed a statistically significant improvement in self-reported stress and anxiety scores among participants who received BeCurie™ compared to their baseline values. A one-way repeated measures ANOVA demonstrated a significant reduction in stress scores (DASS-21 Stress, F(3,18) = 40.77, P<0.001; PSS, F(3, 18) = 7.44, P<0.001) and anxiety scores (DASS-21 Anxiety, F(3, 18) = 51.43, P<0.001; HAM-A, F(3, 18) = 19.89, P<0.001) in the BeCurie™ group compared to baseline. In addition to the improvements in stress and anxiety scores, there was a significant reduction in DASS-21 depression scores (F(3, 18) = 51.30, P<0.001) in the BeCurie™ group compared to their baseline measurements (Table 3, Figure 4). Furthermore, all the subjects were switched to active group after day 30 assessment, to capture the impact of BeCurie™ on converted group. A one-way repeated measures ANOVA demonstrated a significant reduction in stress scores (DASS-21 Stress, F(3, 39) = 21.26, P<0.001; PSS, F(3, 39) = 10.20, P<0.001) and anxiety scores (DASS-21 Anxiety, F(3, 39) = 29.59, P<0.001; HAM-A, F(3, 39) = 12.30, P<0.001) in the treated group. In addition to the improvements in stress and anxiety scores, there was a time dependent improvement in DASS-21 depression scores (F(3, 39) = 21.66, P<0.001) in the BeCurie™ treated subjects (Table 4, Figure 5).

**Figure 4:**
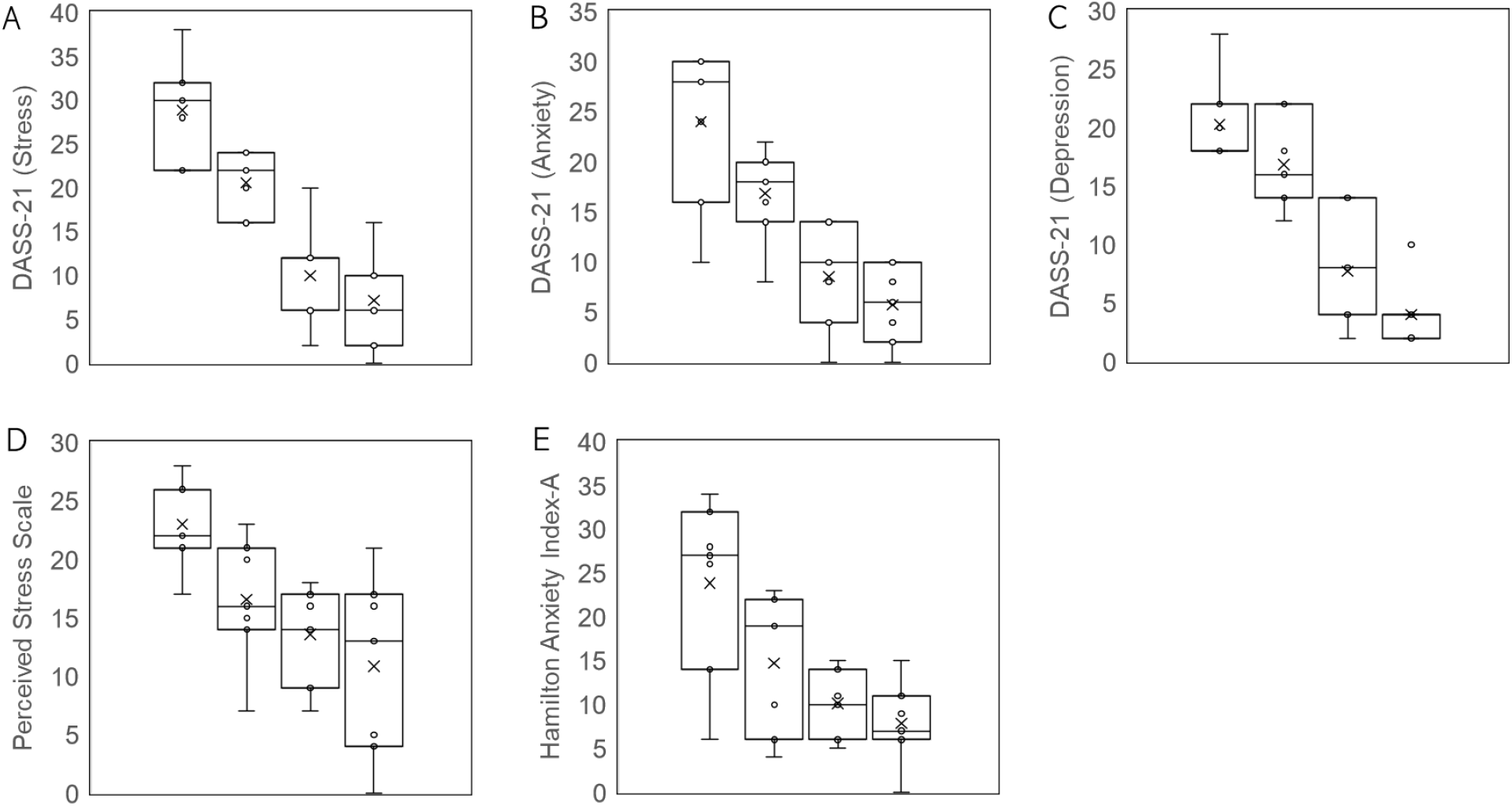
Changes in stress, anxiety, and depression scores in BeCurie™ (n=7) treated group during subsequent analysis on Days 0, 30, 60 & 90. The data is represented in a box chart showing the distribution of data, highlighting the mean and outliers.

**Figure 5:**
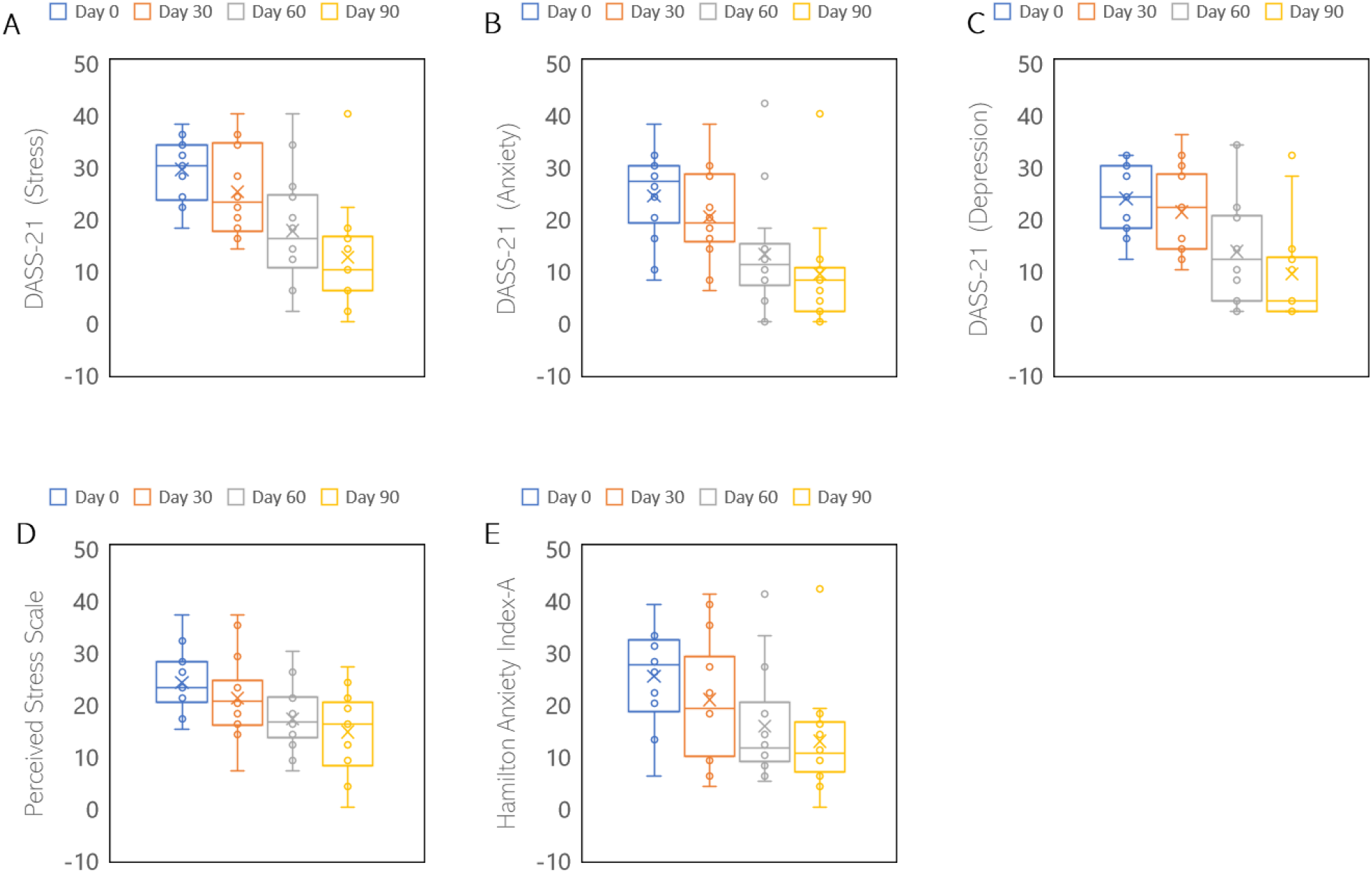
Open Label Subset: Changes in stress, anxiety, and depression scores in BeCurie™ (n=14) treated group during subsequent analysis on Days 0, 30, 60 & 90. The data is represented in a box chart showing the distribution of data, highlighting the mean and outliers.

**Table 3:**
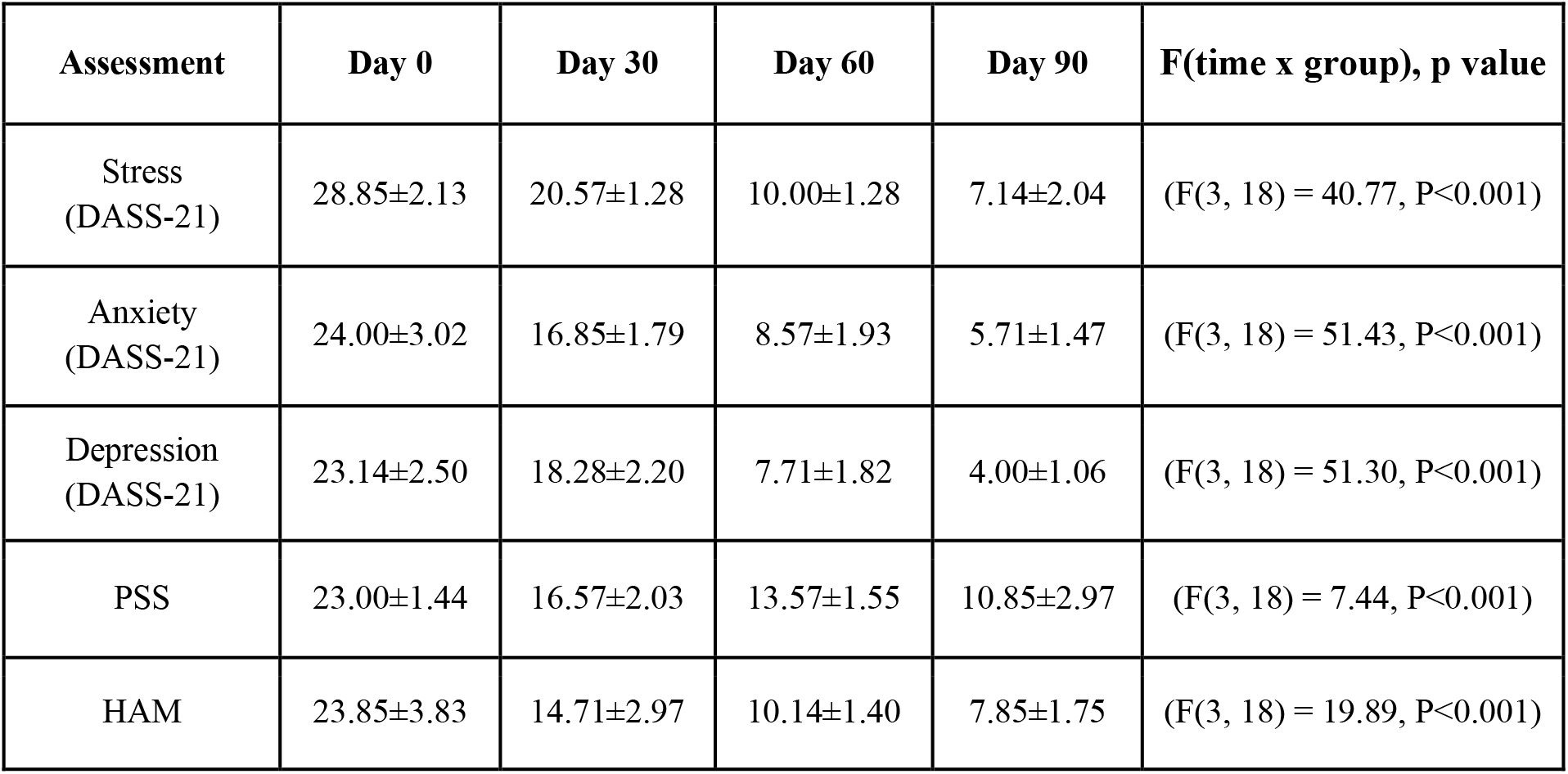
Changes in self-reported stress, anxiety, and depression scores in BeCurie™ treated group (n=7) during subsequent assessments on Day 0, 30, 60 & 90. Values are represented as mean ± SD. *P<0.05 was considered significant. Abbreviations: DASS-21 = The Depression, Anxiety and Stress Scale - 21 Items; PSS = Perceived Stress Scale; HAM-A = The Hamilton Anxiety Rating Scale.

**Table 4:** Open label subset: Changes in self-reported stress, anxiety, and depression scores in BeCurie™ treated group (n=14) during subsequent assessments on Day 0, 30, 60 & 90. Values are represented as mean ± SD. *P<0.05 was considered significant. Abbreviations: DASS-21 = The Depression, Anxiety and Stress Scale - 21 Items; PSS = Perceived Stress Scale; HAM-A = The Hamilton Anxiety Rating Scale.

| Assessment | Day 0 | Day 30 | Day 60 | Day 90 | F(time x group), p value |
| --- | --- | --- | --- | --- | --- |
| Stress (DASS-21) | 29.3±1.6 | 25.0±2.3 | 17.6±2.9 | 12.4±2.6 | (F(3, 39) = 21.26, P<0.001) |
| Anxiety (DASS-21) | 24.3±2.3 | 20.3±2.4 | 13.0±2.9 | 9.3±2.7 | (F(3, 39) = 29.59, P<0.001) |
| Depression (DASS-21) | 23.7±1.7 | 21.1±2.1 | 13.6±2.8 | 9.3±2.6 | (F(3, 39) = 21.66, P<0.001) |
| PSS | 24.1±1.6 | 21.1±2.2 | 17.1±1.7 | 14.5±2.1 | (F(3, 39) = 10.20, P<0.001) |
| HAM | 25.3±2.5 | 20.7±3.1 | 15.6±2.9 | 12.7±2.7 | (F(3, 39) = 12.30, P<0.001). |

### Evaluation of Quality of Life (MQoL) Scores in Subsequent Assessments

The BeCurie™ administered group showed significant improvements in self-reported quality of life scores on Days 30, 60, and 90 when compared to baseline measures. A one-way repeated measures analysis of variance (ANOVA) demonstrated significant improvements in the response to major question items, representing physical, psychological, existential, and support-based wellbeing in the BeCurie™ treated group compared to the placebo group. The respective F-values and P-values are presented in the table (Table 5). Similarly, after cross over, all the subjects showed time dependent improvements in the quality of life parameters in the subsequent assessments on day 60 and day 90 (Table 6).

**Table 5:**
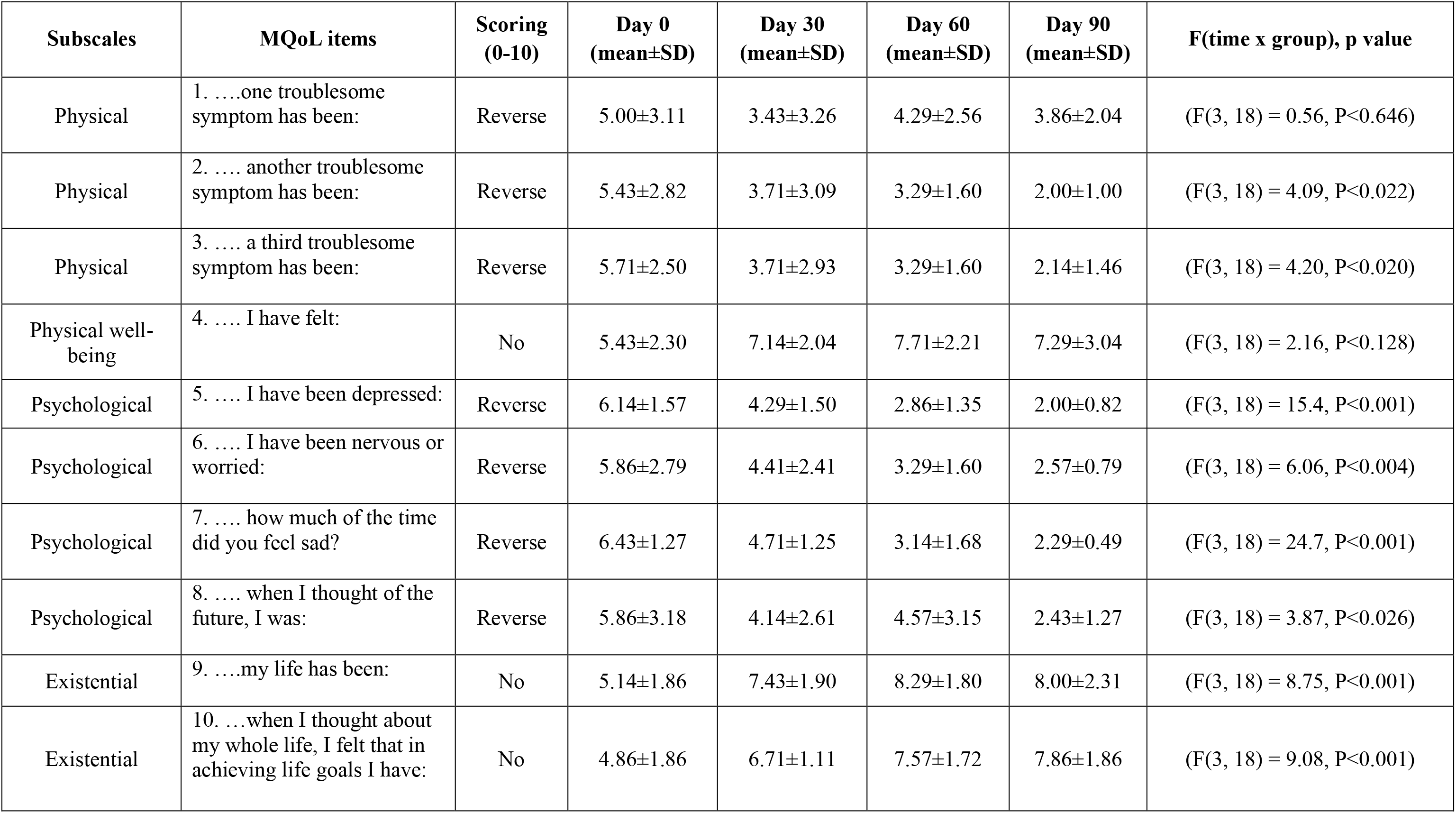
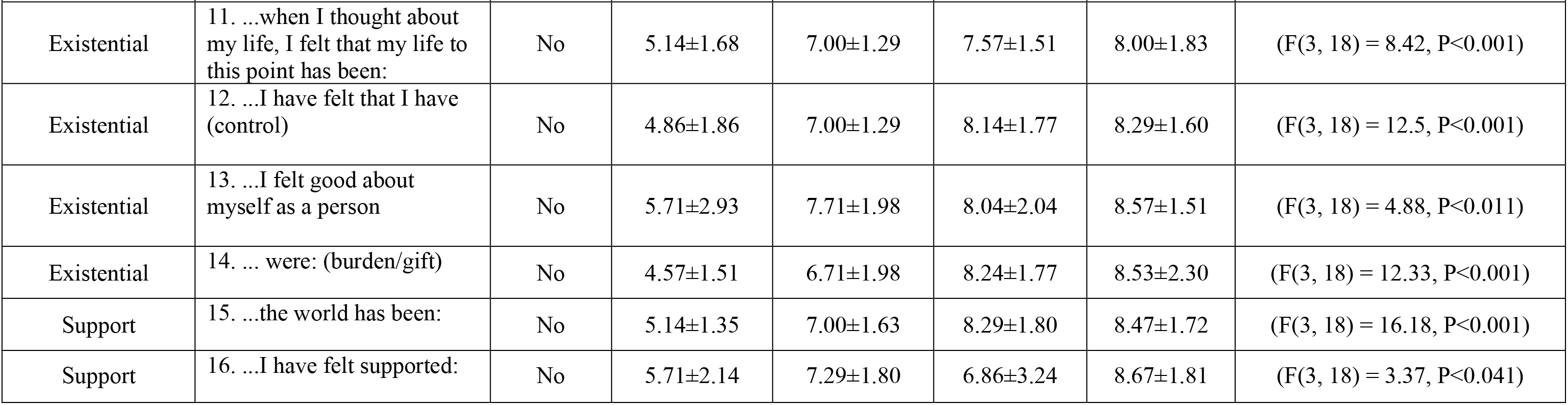
Changes in self-reported Quality of Life scores in BeCurie™ treated arm (n=7) during subsequent assessments on Day 0, 30, 60 & 90. Values are represented as mean ± SD. *P<0.05 was considered significant. Reverse represents the negative scoring (ie., on the scale “10” being poor or feeling dreadful and “0” being good or feeling delighted), No represents positive scoring (ie., on the scale “10” being good or feeling delighted and “0” being poor or feeling dreadful).

**Table 6:**
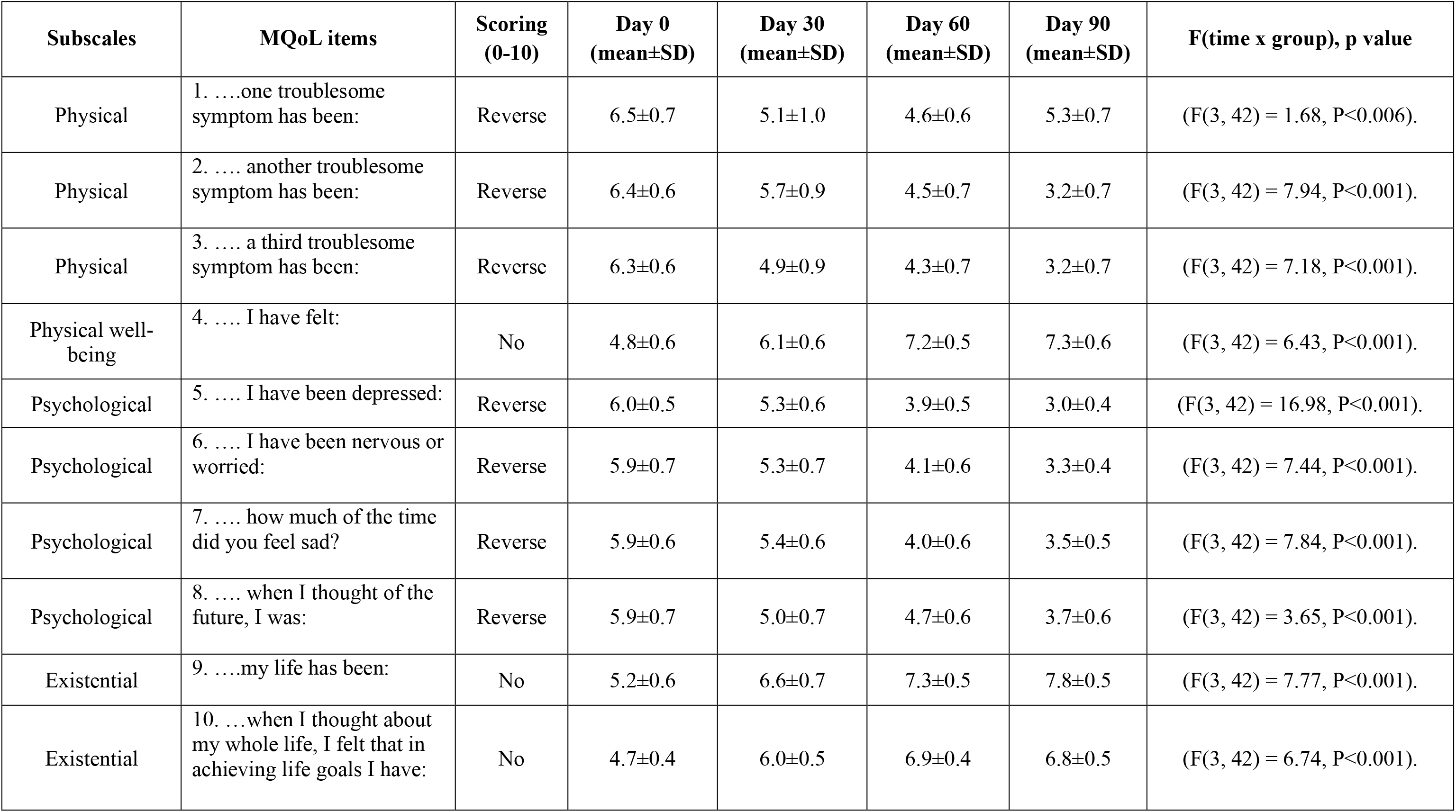
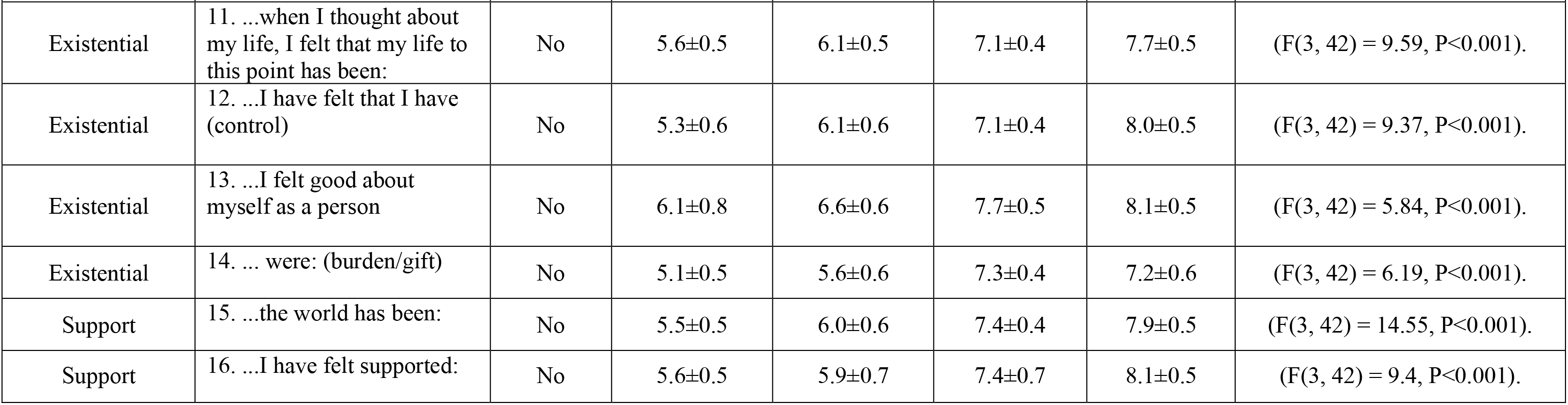
Open label subset: Changes in self-reported Quality of Life scores in BeCurie™ treated subjects (n=14) during subsequent assessments on Day 0, 30, 60 & 90. Values are represented as mean ± SD. *P<0.05 was considered significant. Reverse represents the negative scoring (ie., on the scale “10” being poor or feeling dreadful and “0” being good or feeling delighted), No represents positive scoring (ie., on the scale “10” being good or feeling delighted and “0” being poor or feeling dreadful).

### Clinical Evaluation of Serum Cortisol Levels in Placebo and BeCurie™ Treated Groups

Serum cortisol levels were assessed in both the placebo and BeCurie™ treated groups at baseline (Day 0) and after one month (Day 30) of treatment. There were no significant differences in serum cortisol levels between the placebo and BeCurie™ treated groups at either baseline or Day 30 (Table 7, Figure 6).

**Figure 6:**
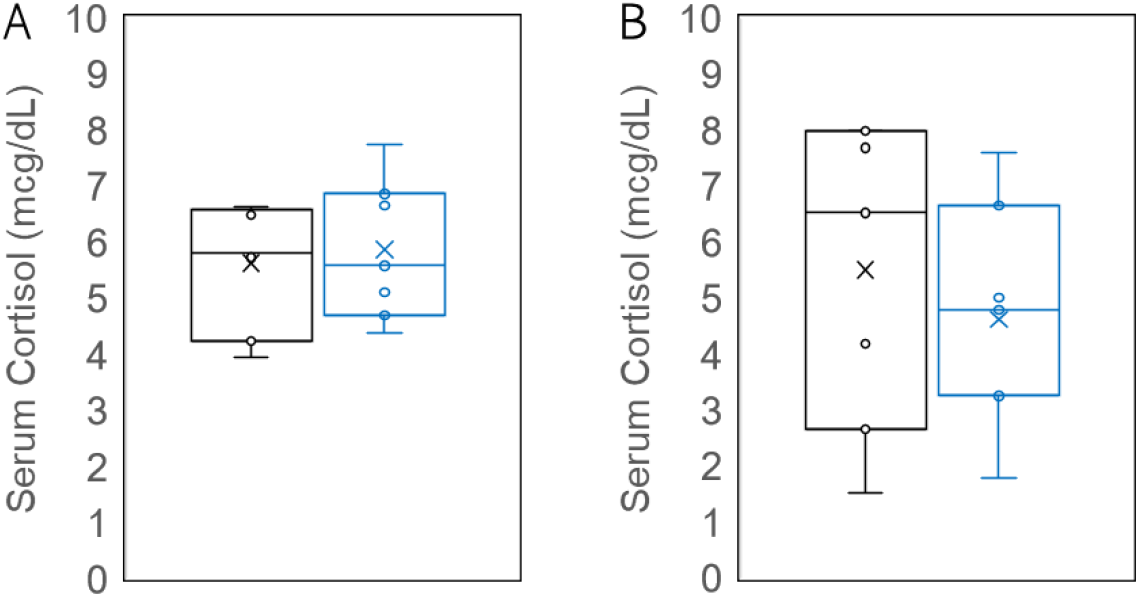
Assessment of serum cortisol levels in A) Placebo (n=7); Day 0 vs Day 30. and B) BeCurie™ (n=7) groups; Day 0 vs Day 30. The data is represented in a box chart showing the distribution of data, highlighting the mean and outliers.

**Table 7:** Changes in serum cortisol levels in Placebo and BeCurie™ arms on Day 0 & Day 30. Values are represented as mean ± SD.

| Group | Serum Cortisol<br>(mcg/dL) |  | 95% CI | t-test | p value |
| --- | --- | --- | --- | --- | --- |
|  | Day 0 | Day 30 |  |  |  |
| **Control**<br>**(Mean±SD)** | 5.63±1.11 | 5.86±1.25 | 1.37 (-1.14, 1.62) | 0.375 | 0.714 |
| **BeCurie™**<br>**(Mean±SD)** | 5.50±2.70 | 4.62±2.04 | 2.79 (-1.91, 3.67) | 0.689 | 0.504 |

### Clinical Evaluation of Blood Parameters in Placebo and BeCurie™ Treated Groups

The safety evaluation of the placebo and BeCurie™ groups was monitored through a comprehensive blood profile assessment on Day 0 and Day 30. No significant changes were observed in any of the assessed blood parameters in either the placebo or BeCurie™ treated groups, and all values remained within normal range. However, some abnormal blood profile levels were identified in both groups independent of treatment. HbA1c levels in the BeCurie™ group exceeded the diabetic threshold (>7; baseline evaluation on Day 0). The placebo group had VLDL values above the threshold (>30 mg/dL), and subjects in both the groups exhibited high triglyceride levels (>150 mg/dL, borderline risk) (Table 8).

**Table 8:**
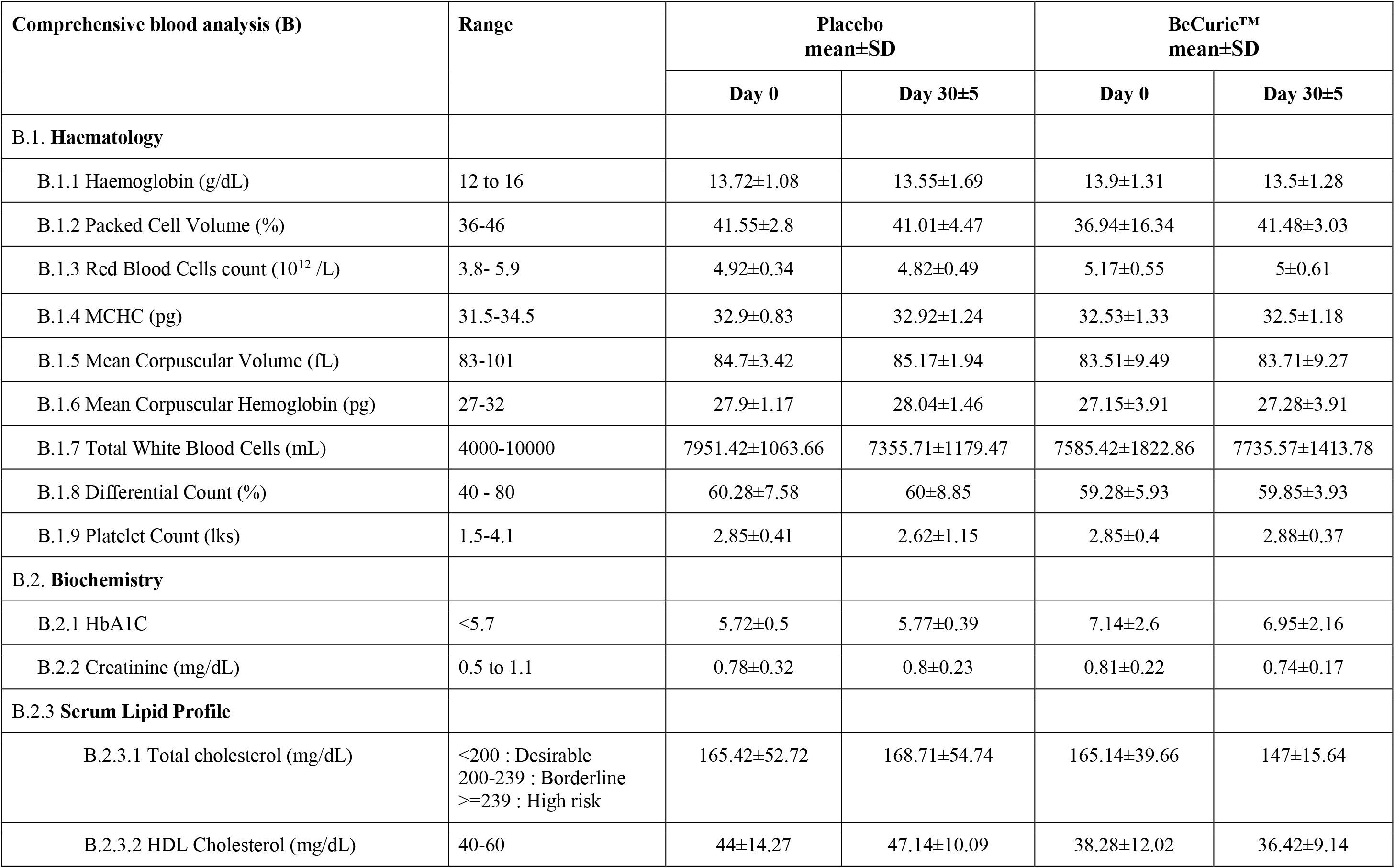
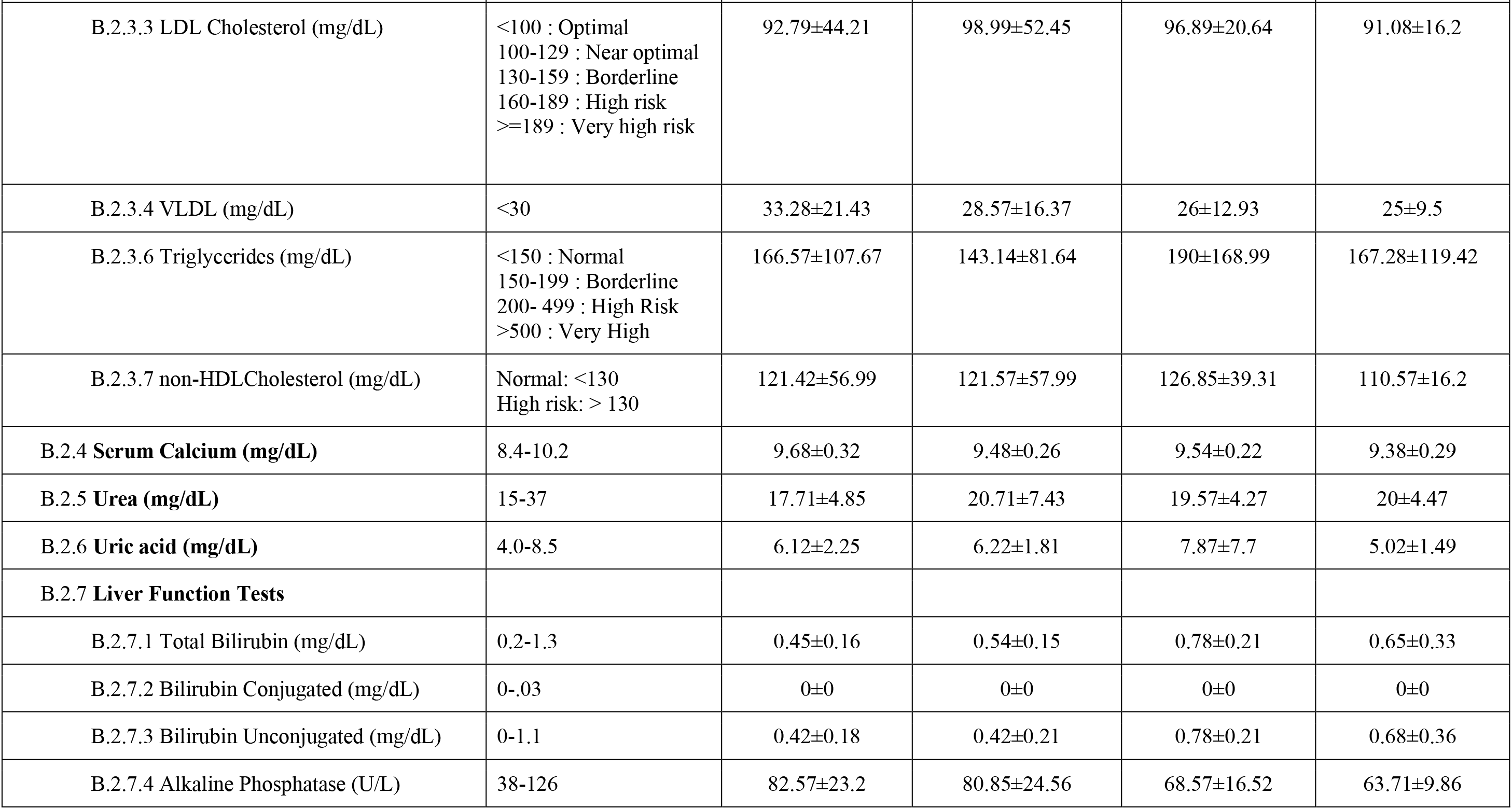
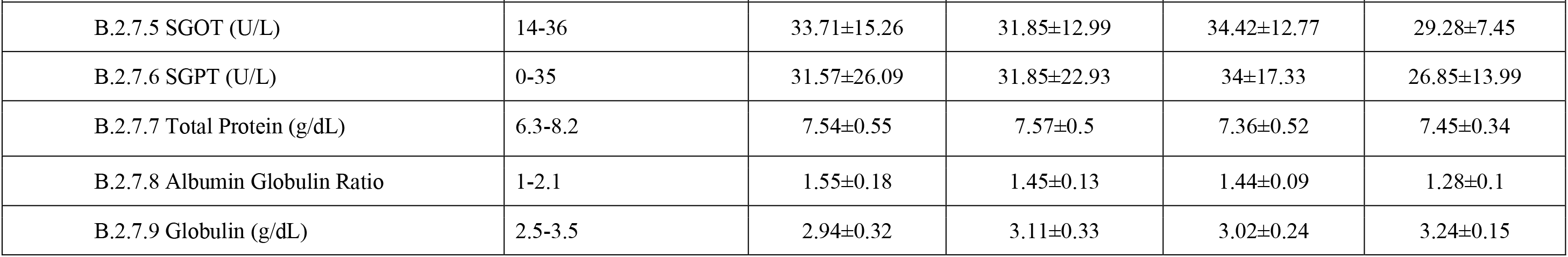
Safety evaluation of complete blood profile in Placebo (n=7) and BeCurie™ (n=7) groups on Day 0 and Day 30. Values are represented as mean ± SD. *P<0.05 was considered significant. Abbreviations: MCHC = Mean Corpuscular Hemoglobin Concentration, HDL = High-density Lipoprotein, LDL = Low-density Lipoprotein, VLDL = Very- Low-density Lipoprotein, SGOT = Serum Glutamic Oxaloacetic Transaminase and SGPT = Serum Glutamate-Pyruvate Transaminase.

## Discussion

The aim of this study was to evaluate the efficacy and safety of VCMF’s generated by the BeCurie™ device in individuals experiencing moderate to severe levels of stress and anxiety. Earlier, Rohan et al, demonstrated the positive impact of the complex weak magnetic fields in elevating mood in patients with bipolar depression^9^. Tsang et al, reported that burst firing of 1 microtesla weak magnetic fields found to be effective for clinical depression, improved mood and vigour compared to the sham-field or other treatments^18^. Persinger et al., demonstrated that application of weak magnetic fields decreases psychometric depression and increases frontal beta activity in normal subjects^32^.

Studies on subjects with severe psychological stress and generalised anxiety disorder (GAD) are identified with increased beta activity in the central part of frontal cortex and decreased alpha rhythm^33–35^. Knyazev et al. reported that trait anxiety and depression were positively correlated with the strength of the reciprocal relationship between alpha and delta oscillations, known as alpha-delta anticorrelation (ADA)^34^. Dadashi et al demonstrated that boosting the amplitude of alpha and theta brain waves in the occipital region can enhance the overall level of functioning and alleviate symptoms of generalized anxiety in individuals with GAD who received treatment^36^. Extremely low-frequency magnetic fields are thought to increase the frequencies of endogenous brain activity of the (2–7 Hz) range towards frequencies of less than or equal to those frequencies of the alpha frequency range (8–13 Hz), in other words, modulation of “neuronal network communication”^37^.

Based on the current evidence^12, 13, 18, 38^, the Becurie™ was developed as a portable, wearable neck device and equipped with latest technical advances to generate VCMF’s within the range of 0.4 to 10 mG. The current study involved eighteen participants, with both males and females enrolled based on DASS-21 eligibility criteria, individuals scoring moderate to severe levels of stress and anxiety. In total fourteen subjects finished the study. Whereas four subjects (three from placebo and one from treatment group) were lost to follow-up assessments (Figure 1).

The primary outcomes of the study indicate the improvements in the self-reported symptoms of stress and anxiety in the BeCurie™ treated group. On Day 30, the BeCurie™ group showed significant reduction in stress and anxiety scores compared to the placebo group. Stress score (DASS-21, 95% CI = 8.6 (0.22, 17.49), p=0.045: PSS, 95% CI = 8.1 (0.89, 17.1), p=0.032) and Anxiety scores (HAM-A, 95% CI=11.9 (0.06, 23.93), p=0.049) were significantly low in the BeCurie™ group when compared to the placebo group (Table 2).

Evaluations of stress, anxiety, and depression scores during subsequent assessments on Days 30, 60, and 90 revealed significant improvements in self-reported stress, anxiety and depression scores in the BeCurie™ treated group. One way repeated measures ANOVA revealed a significant reduction in Stress score (DASS-21 Stress, F(3,18) = 40.77, P<0.001; PSS, F(3, 18) = 7.44, P<0.001) and anxiety score (DASS-21 Anxiety, F(3, 18) = 51.43, P<0.001; HAM-A, F(3, 18) = 19.89, P<0.001) in the BeCurie™ administered group when compared to baseline. Along with the improvements in stress and anxiety scores, there was a significant reduction in DASS-21 depression scores (F(3, 18) = 51.30, P<0.001) when compared to baseline measures in the BeCurie™ administered group (Table 3). In the open label study, we observed a time dependent improvement in the stress, anxiety and depression scores in all the subjects on day 60 and day 90 evaluations (Table 4). Our results are consistent with various other studies that have emphasized the positive impact of weak magnetic fields on various neurological disorders^12, 13, 19–24, 37^. One prominent hypothesis by which VCMF’s may improve neurological stress and anxiety is by modulating alpha frequency in the brain, also known as alpha- event-related desynchronization (alpha-ERD) associated with sensory and cognitive processing of external stimuli including vision, auditory and somatosensory cues^39^.

Furthermore, we noticed a significant improvement in the quality of life (QoL) parameters in the BeCurie™ group when compared to the placebo group (Table 5) and the cross over group (Table 6). The BeCurie™ treatment showed significant improvement in all major domains of QoL, including physical, psychological, existential, and support-based well-being questions. These findings are consistent with the hypothesis that VCMF’s emitted by the BeCurie™ device could have a beneficial effect on mental and emotional well-being, leading to a better overall QoL. The improvements in QoL parameters are in line with the improvements in stress and anxiety measures (Tables 2 to 4).

When an individual faces a stressor that surpasses their current coping mechanisms, the hypothalamic-pituitary-adrenal (HPA) axis is triggered via the chain of events activated between cortex, amygdala, and hippocampus, leading to an elevation in blood cortisol levels^40^. Hence, we assessed the blood cortisol levels in Placebo and BeCurie™ treated groups. However, we did not observe any significant changes in cortisol levels between the placebo and BeCurie™ groups (Table 7 & Figure 6), indicating that VCMF’s generated by BeCurie™ did not have any effect on the hypothalamic-pituitary-adrenal (HPA) axis.

Weak magnetic fields are proven to be extremely safe for long term use without any side effects or adverse effects. Moreover, the variable complex weak magnetic fields generated by the BeCurie™ device are ∼10 and 10^6^-fold lower in magnetic field strength when compared to geomagnetic fields (30 to 70 µT) and TMS devices (1-3 Tesla)^41–44^. No adverse events were reported during the entire duration of the study in both the placebo and BeCurie™ treated subjects. Comprehensive blood profile assessment showed no significant changes or signs of toxicity in either the placebo or BeCurie™ groups, indicating that VCMF’s generated by BeCurie™ doesn’t adversely impact physiological parameters (Table 8). However, it is worth noting that a few abnormal blood profile levels were identified in both the placebo and BeCurie™ groups independent of treatment. HbA1c levels in the BeCurie™ group were found to be high (>7, diabetic, baseline evaluation on day 0). Very low-density lipoprotein (VLDL) values were above the threshold in the placebo group (>30 mg/dL), and the subjects enrolled in both the cohorts were found to have high triglyceride levels (>150 mg/dL, borderline risk) (Table 8).

While our results demonstrate preliminary evidence for the efficacy and safety of VCMF’s therapy in managing self-perceived levels of stress and anxiety, there are few limitations to our study. Mechanisms by which VCMF’s generated by BeCurie™ device modulate the neuronal activity is yet to be understood, one such study by potentially evaluating the changes in regional brain frequencies upon BeCurie™ therapy in subjects with stress and anxiety might provide some insights into its neuromodulatory activity^45, 46^. One of the major limitations is the small sample size, which may affect the generalizability of the findings. Another limitation is the lack of a follow-up assessment beyond 90 days, which could have provided insights into the long-term effects of BeCurie™ on stress and anxiety symptoms.

## Conclusion

Notwithstanding the limitations, this study provides preliminary evidence that VCMF’s generated by the BeCurie™ device can be a potential non-invasive intervention for managing stress and anxiety. Our findings provide preliminary evidence for the efficacy and safety of BeCurie™ and warrant further investigations in larger randomized placebo led trials with longer follow-up periods and more diverse populations to confirm our findings.

## Data Availability

All data produced in the present study are available upon reasonable request to the authors

## Acknowledgements

The authors thank Mr Swamy Chikurumelli (study conduct), Dr Srinivas Bhandaru (study management), Mr Lakshman (study coordinator) for their support with the conduct of the clinical study and thank the management of Yashoda hospitals for resource management and their clinical collaboration.

## Competing Interest Statement

All authors have completed the ICMJE uniform disclosure form at www.icmje.org/coi_disclosure.pdf and declare: Mr Shyam Sunder Pasumarthi and Dr Lalitha Palle are the founders of Aether Mindtech Solutions Pvt Ltd and sponsored the study and has commercial interest over the device. Dr Chaitanya Chakravarthi Gali is employed by My Pura Vida Wellness Pvt Ltd and has no competing interest. Aether Mindtech Solutions Pvt Ltd and My Pura Vida Wellness Pvt Ltd had no role in the planning, conducting and analysis of data of this randomized controlled trial. Dr Mohan Krishna Jonnalagadda declares that he has no support from any organization for the submitted work; no financial relationships with any organizations that might have an interest in the submitted work in the previous three years; no other relationship or activities that could appear to have influenced the submitted work.

## Notes

### Clinical Trial

CTRI/2022/03/041445

### Clinical Protocols

https://ctri.nic.in/Clinicaltrials/showallp.php?mid1=65703&EncHid=&userName=becurie

### Funding Statement

The study was funded by the sponsor Aether Mindtech Solutions Private Limited.

### Author Declarations

The study was approved by the Institutional Ethics Committee of Yashoda Academy of Medical Education and Research (IEC-YAMER) with registration number ECR/49/Inst/AP/2013/RR-19.

